# Identification of REM Sleep Behavior Disorder by Magnetic Resonance Imaging and Machine Learning

**DOI:** 10.1101/2021.09.18.21263779

**Authors:** Jie Mei, Shady Rahayel, Christian Desrosiers, Ronald B. Postuma, Jacques Montplaisir, Julie Carrier, Oury Monchi, Johannes Frasnelli, Jean-François Gagnon

**Author notes:** These authors contributed equally to this work (Jie Mei, Shady Rahayel). These authors share senior authorship (Johannes Frasnelli, Jean-François Gagnon). Current affiliation: 1. The Brain and Mind Institute, University of Western Ontario, London, ON, Canada. 2. Department of Computer Science, University of Western Ontario, London, ON, Canada. **Contact information:** Dr. Jean-François Gagnon, Département de psychologie, Université du Québec à Montréal C.P. 8888 succ. Centre-ville, Montréal (Québec), H3C 3P8, Canada. **Funding sources:** The Canadian Institutes of Health Research (CIHR), the Fonds de recherche du Québec – Santé (FRQ-S), the Réseau intersectoriel de recherche en santé de l’Université du Québec (RISUQ), the Centre de recherche en neurosciences cognitives de l’Université du Québec à Montréal (NeuroQAM), Parkinson Canada, and the W. Garfield Weston Foundation.

## Abstract

**Background:** Idiopathic rapid eye movement sleep behavior disorder (iRBD) is a major risk factor for synucleinopathies, and patients often present with clinical signs and morphological brain changes. However, there is a heterogeneity in the presentation and progression of these alterations, and brain regions that are more vulnerable to neurodegeneration remain to be determined.

**Objectives:** To assess the feasibility of morphology-based machine learning in the identification and subtyping of iRBD.

**Methods:** For the classification tasks [iRBD (n=48) vs controls (n=41); iRBD vs Parkinson’s disease (n=29); iRBD with mild cognitive impairment (n=16) vs without mild cognitive impairment (n=32)], machine learning models were trained with morphometric measurements (thickness, surface area, volume, and deformation) extracted from T1-weighted structural magnetic resonance imaging. Model performance and the most discriminative brain regions were analyzed and identified.

**Results:** A high accuracy was reported for iRBD vs controls (79.6%, deformation of the caudal middle frontal gyrus and putamen, thinning of the superior frontal gyrus, and reduced volume of the inferior parietal cortex and insula), iRBD vs Parkinson’s disease (82%, smaller volume and surface area of the insula, lower thinning of the entorhinal cortex and lingual gyrus, and greater volume of the fusiform gyrus), and iRBD with vs without mild cognitive impairment (84.8%, thinning of the pars triangularis, superior temporal gyrus, transverse temporal cortex, larger surface area of the superior temporal gyrus, and deformation of isthmus of the cingulate gyrus).

**Conclusions:** Morphology-based machine learning approaches may allow for detection and subtyping of iRBD, potentially enabling efficient preclinical identification of synucleinopathies.

## Introduction

Isolated/idiopathic rapid eye movement (REM) sleep behavior disorder (iRBD) is a parasomnia characterized by loss of muscle atonia and abnormal motor manifestations during REM sleep (1). iRBD is a prodromal alpha-synucleinopathy, with most patients developing Parkinson’s disease (PD) or dementia with Lewy bodies (DLB) over time (2). iRBD patients already show clinical and brain changes that are reminiscent of overt synucleinopathies (3–5), including cognitive impairment and brain atrophy (1,4–7). However, the identification and progression of these changes vary widely in the iRBD population (4–7). In particular, mild cognitive impairment (MCI), present in about one third of iRBD patients (5,6), is associated with a higher likelihood of phenoconversion to a dementia-compared to a parkinsonism-first phenotype (5,6,8).

Studies using structural magnetic resonance imaging (MRI) in iRBD reported extensive cortical and subcortical changes (4,5,7,9–14), which are more pronounced in patients with MCI (5,15). Findings have included frontal, temporal and occipital cortical thinning (9–11), subcortical shape contraction in the basal ganglia and hippocampus (10,12), and gray matter volume alterations in the prefrontal cortex, caudate, brainstem, cerebellum, and parahippocampal gyrus (9,13,14). Moreover, a brain volume deformation signature that could predict development of DLB at the individual level has been identified in iRBD (16). However, most of these studies have only looked at morphological changes one metric at a time, thereby preventing identifying sets of regions that may discriminate between iRBD patients and healthy individuals (controls) or between iRBD subtypes (presence of MCI), while taking into consideration several different morphological features that can be derived from every brain region.

Machine learning have been increasingly applied to the identification and prognosis of neurodegenerative diseases including PD and DLB (17–20). Few studies have used machine learning for the identification of iRBD with polysomnogram (PSG), electroencephalogram (EEG), motor, and olfaction measures (21–25). To our knowledge, only one study has applied machine learning to diffusion tensor imaging for the identification of iRBD vs controls, and achieved an accuracy of 87.5% (26). However, this study has a small sample size and no distinction based on the cognitive profile. Given the ability of brain changes, mainly those associated with gray matter alterations, to identify MCI and predict phenoconversion in iRBD (15,16), an assessment of morphometric changes, along with a thorough evaluation of the discriminative power of single versus multiple brain region(s) in the identification and subtyping of iRBD, is of relevance. Moreover, machine learning has been applied to the extraction of relevant pre-clinical and clinical features, and can be used in inter-modality registration of different types of data (17,27), potentially enabling improved identification of iRBD.

In this study, we derived regional brain morphological measurements including cortical thickness, volume, surface area and tissue deformation to assess whether machine learning models differentiate between **(a)** iRBD patients and controls, **(b)** iRBD and PD patients, and **(c)** iRBD patients with and without MCI. We hypothesized brain morphology can differentiate between controls, PD patients and iRBD subtypes with high accuracy.

## Methods

### Participants

iRBD patients were enrolled at the Centre for Advanced Research in Sleep Medicine of the *Centre Intégré universitaire de santé et de services sociaux du Nord-de-l’Île-de-Montréal* – *Hôpital du Sacré-Cœur de Montréal (CIUSS-NÎM-HSCM)*. All patients met the diagnostic criteria for iRBD based on the International Classification of Sleep Disorders, Third Edition and PSG (28,29). iRBD patients were excluded if they presented with parkinsonism or dementia (30,31). A history of brain injury, head trauma, stroke, claustrophobia, EEG abnormalities suggesting epilepsy, encephalitis, or other neurological disorders also led to exclusion. Controls without PD, iRBD, and MCI were recruited through newspaper advertisements or by word of mouth. They were subjected to the same exclusion criteria as iRBD patients. All participants were part of previous studies on neuroimaging in iRBD (9,12,15,16,32).

PD patients were recruited from the Department of Neurology of the Montreal General Hospital and the *Unité des troubles du mouvement André-Barbeau* of the *Centre Hospitalier de l’Université de Montréal*. Inclusion criteria were: 1) a diagnosis of idiopathic PD as the likeliest cause (30), 2) being 45-85 years old, 3) disease duration ≤10 years, 4) Hoehn & Yahr stage ≤3, 5) absence of dementia or any major psychiatric disorder, 6) respiratory event index ≤20, and 7) absence of a history of head injury, stroke, brain tumor, cerebrovascular disease, and chronic obstructive pulmonary disease or abnormal EEG features suggesting epilepsy. These patients were part of previous studies on neuroimaging in PD (16,33).

All participants were part of research protocols approved by local ethics committees (CIUSSS-NÎM-HSCM) and CIUSSS *du Centre-Sud-de-l’Île-de-Montréal* – *Comité d’éthique de la recherche vieillissement-neuroimagerie,* Montreal, Canada) and provided written informed consent.

### Neuropsychological assessment

Participants underwent neurological and neuropsychological assessments including the Unified Parkinson’s Disease Rating Scale part III (34), and, in iRBD patients only, the Montreal Cognitive assessment (MoCA) for cognitive screening (35). MCI was diagnosed with the neuropsychological assessment and a consensus between the neurologist and neuropsychologist based on the following criteria: 1) subjective cognitive complaints by the patient, the spouse or the caregiver, as measured using the Cognitive Failures Questionnaire (36) or as assessed during the semi-structured interview, 2) the presence of objective cognitive impairment, as defined by a performance score at least 1.5 SD below the standardized mean on at least 2 tasks within a single cognitive domain, namely attention/executive functions, learning and verbal memory, or visuospatial abilities, 3) preserved daily life functioning, 4) no dementia, and 5) cognitive deficits not being explained solely by a medication or another medical condition (8,15).

### MRI

#### Acquisition

T1-weighted scans were acquired in all participants using a 3T Siemens TrioTIM scanner with a 12-channel head matrix coil and an MP-RAGE sequence (parameters: repetition time: 2.3s, echo time: 2.91ms, inversion time: 900ms, flip angle: 9°, field of view: 256×240mm, matrix resolution: 256×240mm, voxel size: 1×1×1mm, bandwidth: 240Hz/Px, 160 slices).

#### Morphological processing

Cortical surface processing was first conducted using FreeSurfer, version 6.0.0 (37,38). Processing steps included non-brain tissue removal, Talairach transformation, segmentation of subcortical white matter and gray matter subcortical volumetric structures, intensity normalization, and cortical reconstruction. Quality control was performed visually at a slice level and pial and white matter surface errors were corrected manually by a trained rater (S.R.). Cortical thickness, surface area, and volume maps were then parcellated into 34 cortical regions per hemisphere using the Desikan-Killiany atlas (39). Regional surface area and volume values were normalized by the estimated total intracranial volume since these measurements scale with head size.

Deformation-based morphometry (DBM) processing was applied to the T1-weighted images using CAT12 (http://www.neuro.uni-jena.de/cat/) to generate gray matter tissue deformation maps. DBM quantified volume differences by computing the deformations needed to perform nonlinear transformation of an individual brain to a template space (40). The Jacobian determinant maps were smoothed using a 12-mm FWHM kernel and used as a marker of local brain deformation. Maps were parcellated into 83 regions, namely 34 cortical and 7 subcortical regions (putamen, caudate, pallidum, thalamus, hippocampus, amygdala, accumbens area) for each hemisphere plus the brainstem, according to the Desikan-Killiany atlas (39). The deformation value of each region represented the mean deformation of all voxels assigned to the region.

### Feature selection and machine learning

We first performed feature selection to identify morphometric measurements with high discriminative power. To ensure no data were used both for feature selection and model validation, the dataset was split into a training set and a test set in a stratified manner. To understand whether feature selection and model performance were affected by patient distribution of the train-test split, splitting was performed for 25 times using a train:test split ratio of 9:1 (for iRBD vs controls and iRBD vs PD) or 4:1 (for iRBD with MCI vs iRBD without MCI), to generate 25 unique splits for obtaining the averaged model performance (Supplementary Figure 1). Given the number of features (68, 68, 68, and 83 for thickness, surface area, volume and deformation respectively), we used a random forest classifier (n_estimators = 10) on the training set with 5-fold cross-validation to select features based on relative importance averaged over 1,000 iterations.

For the discrimination between iRBD and control, machine learning models were trained with 1, 2, 3, or 5 out of the 15-20 most relevant brain regions for each individual modality. To examine whether combining different morphometric modalities leads to higher model performance, we performed the same procedures after combining regional measurements across modalities (n=287). For iRBD vs PD patients and iRBD patients with MCI vs without MCI, we merged across all modalities for model training without examining classification performance of individual modalities.

We used an expanded grid-search with 5-fold cross-validation to search through models including: 1) logistic regression, 2) decision tree, 3) random forest, 4) k-nearest neighbors (KNN), 5) support vector machine (SVM), and 6) boosting models, and their hyperparameters. All morphometric measurements were centered to the mean and scaled to unit variance. All procedures were implemented using Scikit-learn, version 0.17 (41).

### Model evaluation and statistical analysis

To identify brain regions showing atrophy, we performed Mann-Whitney U tests followed by the Benjamini-Hochberg procedure for controlling false discovery for multiple comparisons (42). Given the imbalanced dataset, in addition to accuracy, metrics including recall, precision, and F1-score were used. Recall, also known as sensitivity or true positive rate, is defined as the number of true positives divided by the sum of true positives and false negatives. Precision is defined as the number of true positives divided by the sum of true positives and false positives. The F1-score is the harmonic mean of precision and recall, and accuracy indicates the ratio of number of correct predictions to number of all predictions. Data are shown as mean (SD). Statistical analyses were performed using pandas, version 0.25.3 (43) and scikit-posthocs, version 0.6.7 (44).

## Results

### Demographics and clinical data

Of the 59 iRBD patients recruited, 11 were excluded for parkinsonism or dementia at presentation or unclear clinical presentation/borderline electromyogram thresholds for REM sleep muscle atonia. All participants survived quality control except for one PD patient who had abnormal surface reconstruction in the occipital lobe, yielding 48 iRBD patients, 29 PD patients [mean (SD), PD duration since diagnosis: 3.8 (2.7) years; levodopa equivalent dosage: 536.5 (283.1) mg/daily; Hoehn & Yahr stage: 2.2 (0.8); 41% (12/29) with MCI; 48% (14/29) with RBD], and 41 controls (Table 1) for MRI processing. Sixteen (33%) iRBD patients had MCI. There were no significant differences in age and education between the groups. The proportion of men was higher in iRBD than PD patients. PD had a higher UPDRS-III total score than iRBD patients. Moreover, iRBD patients with MCI performed poorer than those without MCI on the MoCA.

**Table 1.** Demographic and clinical features in participants.

| Variable | iRBD<br>(A) | iRBD<br>with MCI<br>(B) | iRBD<br>without MCI<br>(C) | PD<br>(D) | Controls<br>(E) | p value,<br>A vs. E | p value,<br>A vs. D | p value,<br>B vs. C |
| --- | --- | --- | --- | --- | --- | --- | --- | --- |
| Age at MRI, years | 65.8 (6.4) | 67.9 (4.6) | 64.8 (7.0) | 64.8 (8.3) | 63.2 (8.2) | 0.09<sup>b</sup> | 0.56<sup>b</sup> | 0.080<sup>b</sup> |
| Men, n (%) | 37 (77) | 10 (63) | 27 (84) | 15 (50) | 25 (61) | 1.00<sup>c</sup> | 0.014<sup>c</sup> | 0.089<sup>c</sup> |
| Education, years | 13.3 (3.7) | 11.9 (3.8) | 14.0 (3.4) | 14.9 (3.8) | 14.6 (4.1) | 0.18<sup>b</sup> | 0.061<sup>b</sup> | 0.058<sup>b</sup> |
| RBD duration since<br>symptom onset, years | 12.1 (12.2) | 13.3 (13.4) | 11.5 (11.7) | - | - | - | - | 0.47<sup>d</sup> |
| UPDRS-III, total score<sup>a</sup> | 4.3 (3.6) | 5.6 (5.0) | 3.7 (2.6) | 20.6 (9.2) | - | - | <0.001<sup>d</sup> | 0.24<sup>d</sup> |
| MoCA, total score | 25.7 (2.8) | 24.1 (3.3) | 26.9 (1.8) | - | - | - | - | 0.005<sup>d</sup> |

Values are presented as mean (SD).
PD: Parkinson's disease, iRBD: idiopathic rapid eye movement sleep behavior disorder, MCI: mild cognitive impairment, UPDRS-III: Unified Parkinson's Disease Rating Scale part III, MoCA: Montreal Cognitive Assessment.
<sup>a</sup>The UPDRS-III was measured in the 'on' state in PD patients.
<sup>b</sup>Student t test.
<sup>c</sup>Chi-squared test.
<sup>d</sup>Mann-Whitney U test.

### Morphometric measurements

Of the 287 morphometric features, 37 (12.9%) significantly differed between iRBD patients and controls, 20 (7.0%) between iRBD and PD patients, and 27 (9.4%) between iRBD patients with or without MCI (Supplementary Tables 1, 4 and 5). Compared to controls, iRBD patients had cortical thinning, reduced volume, and tissue deformation in several brain regions (Supplementary Table 1). Compared to PD, iRBD patients had mixed results for cortical thickness, but with reduced volume in PD (bilateral fusiform gyrus) and tissue deformation, mainly in left subcortical regions, in iRBD (Supplementary Table 4). Compared to patients without MCI, those with MCI had cortical thinning, larger surface area, and tissue deformation, mainly in posterior regions (Supplementary Table 5).

### Feature importance and machine learning-based classification

Although 5 brain regions that contributed to the highest overall model performance were reported, it should be noted that in all classification tasks, feature importance of the most relevant brain regions (top 5%-10%) were often comparable (Supplementary Tables 2 and 3) and could lead to classification performance that was only 2%-5% lower than the reported results.

#### iRBD vs Controls

The highest accuracy (79.6%) was obtained when measurements of all morphometric modalities were merged to select 5 features for model training, namely deformation of the left caudal middle frontal gyrus and right putamen, thinning of the left superior frontal gyrus, and reduced volume of the right inferior parietal cortex and insula (Table 2 and Figures 1a-e and 2). When using single modalities for model training, a classification accuracy of 79.1% was achieved with 5 features selected from DBM-based measurements, namely tissue deformation in the left caudal middle frontal gyrus, paracentral lobule, thalamus, and bilateral putamen. When training the model with 1 feature derived from DBM or across all modalities, deformation of the left caudal middle frontal gyrus led to an accuracy of 71.1%.

**Figure 1.**
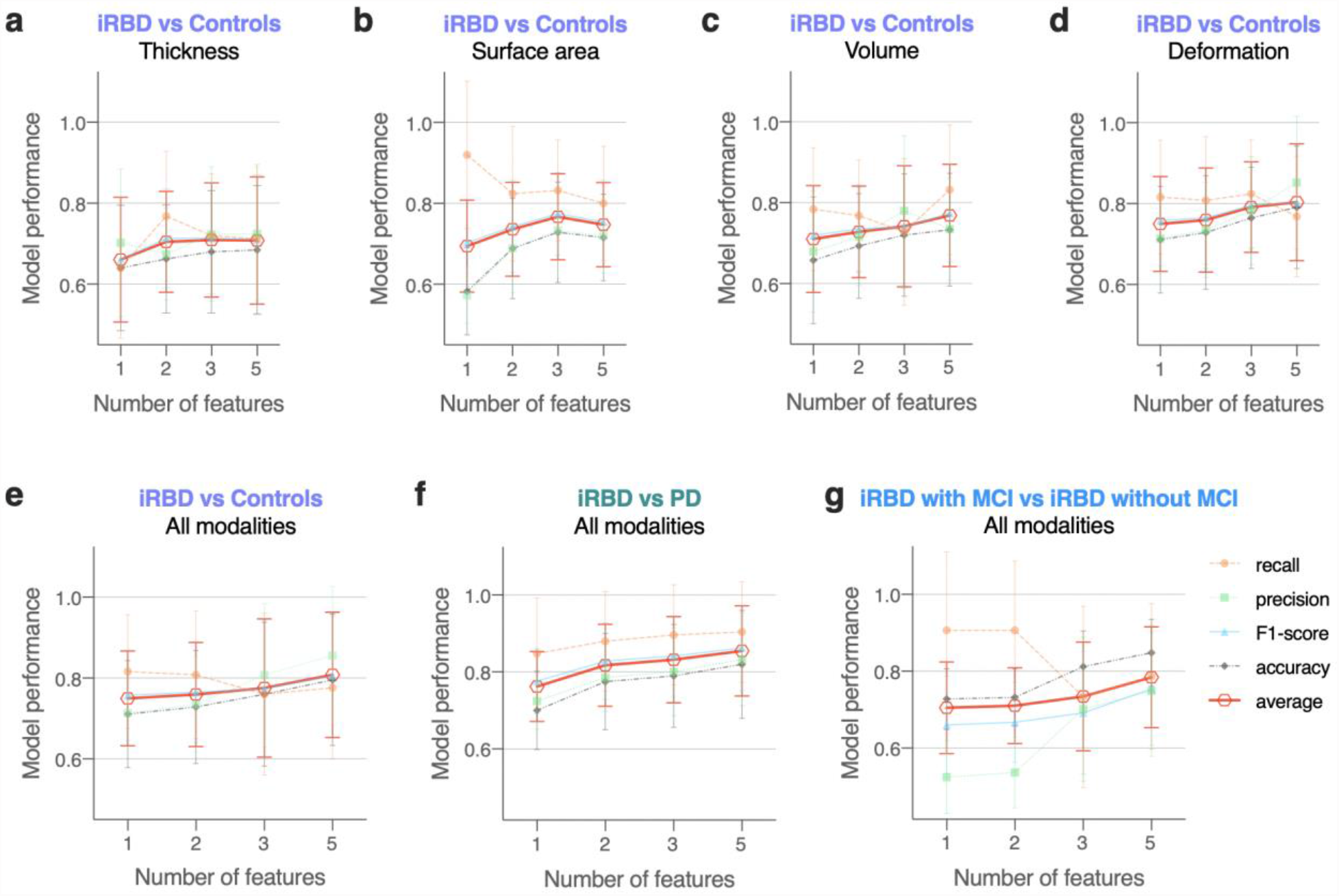
Results of machine learning models with the highest overall performance as measured by recall, precision, F1-score, accuracy, and average of the four metrics. (a-d) Outcomes of individual morphometric modalities in the differentiation between iRBD patients and controls. (e-g) Outcomes when all four morphometric modalities were merged and used in model training, for iRBD patients vs Controls (e), iRBD vs PD patients (f) and iRBD patients with MCI vs without MCI (g). iRBD: idiopathic rapid eye movement sleep behavior disorder, PD: Parkinson’s disease, MCI: mild cognitive impairment. Error bars indicate standard deviation.

**Figure 2.**
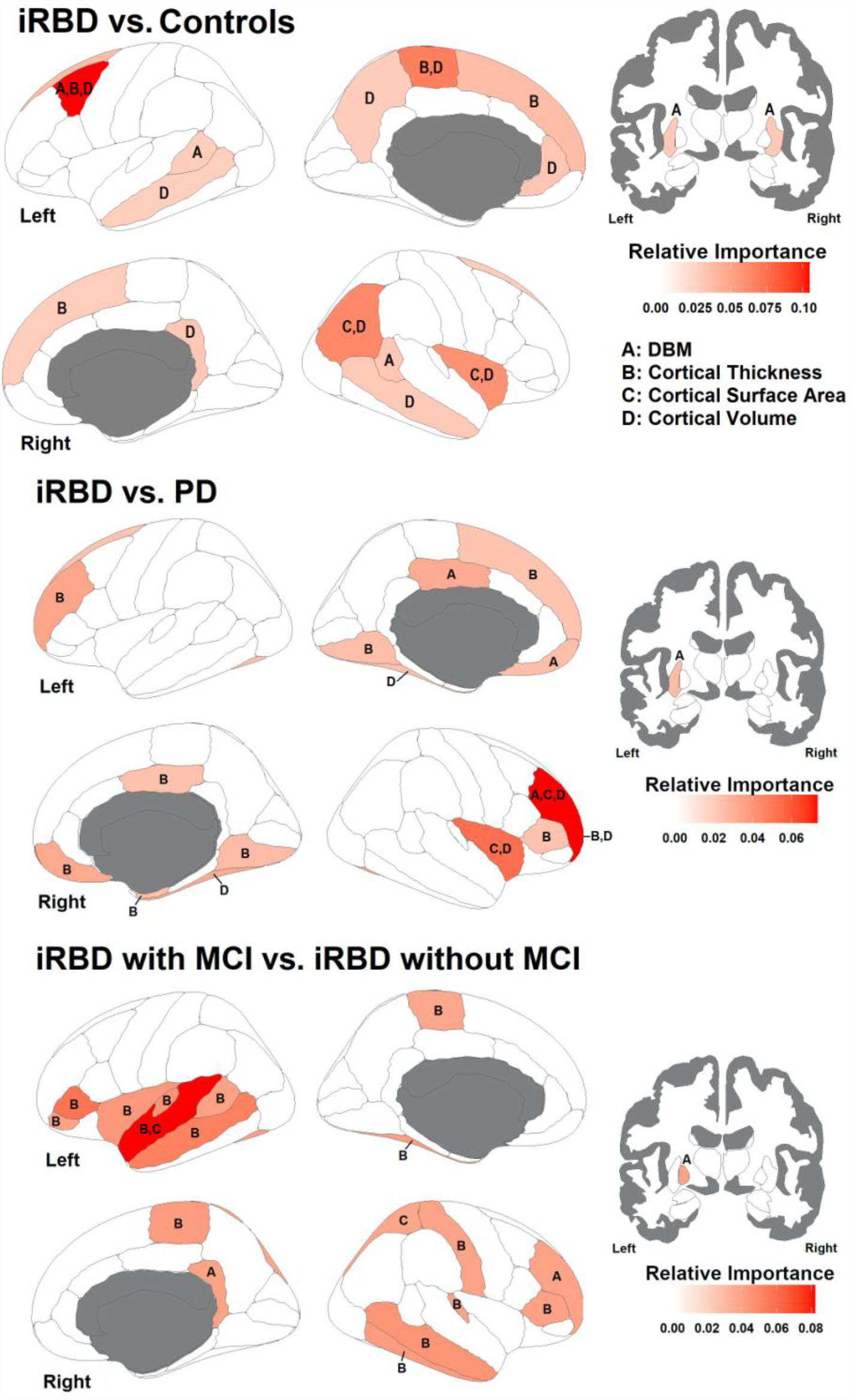
Relative importance of the top 20 most relevant brain regions in the discrimination between iRBD vs Controls (top), iRBD vs PD patients (middle) and iRBD patients with vs without MCI (bottom). The white-red color bar represents the relative importance of the feature in discriminating groups, with redder regions representing a more relevant feature. Every region from the top 20 is labeled from A-D in terms of the contributing structural metric (legend on the right). DBM: deformation-based morphometry, iRBD: idiopathic rapid eye movement sleep behavior disorder, MCI: mild cognitive impairment, PD: Parkinson’s disease.

**Table 2.** Results of the best performing models, brain regions and associated morphometric measurements used in the differentiation between participants.

| Modality | Number of features | Recall | Precision | F1-score | Accuracy | Machine learning model | Brain region and characteristics in iRBD vs controls |
| --- | --- | --- | --- | --- | --- | --- | --- |
| Cortical thickness | 1 | 0.640<br>(0.173) | 0.702<br>(0.182) | 0.661<br>(0.153) | 0.640<br>(0.155) | Linear SVM, C = 1 | thinning of paracentral lobule in iRBD (lh) |
|  | 5 | 0.712<br>(0.183) | 0.725<br>(0.166) | 0.710<br>(0.160) | 0.684<br>(0.159) | Linear SVM, C = 1 | thinning of paracentral lobule in iRBD (lh)<br>thinning of medial orbital frontal cortex in iRBD (lh)<br>thicker banks of the superior temporal sulcus in iRBD (lh)<br>thinning of temporal pole in iRBD (lh)<br>thinning of frontal pole in iRBD (rh) |
| Surface area | 1 | 0.920<br>(0.183) | 0.573<br>(0.069) | 0.703<br>(0.108) | 0.582<br>(0.108) | Linear SVM, C = 1 | larger surface area of isthmus of cingulate cortex in iRBD (lh) |
|  | 5 | 0.800<br>(0.141) | 0.721<br>(0.093) | 0.754<br>(0.103) | 0.716<br>(0.107) | Linear SVM, C = 1 | larger surface area of isthmus of cingulate cortex in iRBD (lh)<br>reduced surface area of rostral anterior cingulate cortex in iRBD (lh)<br>larger surface area of precentral gyrus in iRBD (lh)<br>reduced surface area of rostral middle frontal gyrus in iRBD (lh)<br>larger surface area of temporal pole in iRBD (lh) |
| Volume | 1 | 0.784<br>(0.152) | 0.680<br>(0.150) | 0.720<br>(0.124) | 0.658<br>(0.157) | SVM with RBF kernel, C = 1 | reduced volume of caudal middle frontal gyrus in iRBD (lh) |
|  | 5 | 0.832<br>(0.160) | 0.735<br>(0.126) | 0.775<br>(0.121) | 0.733<br>(0.140) | SVM with RBF kernel, C = 1 | reduced volume of caudal middle frontal gyrus in iRBD (lh)<br>reduced volume of paracentral lobule in iRBD (lh)<br>reduced volume of rostral anterior cingulate cortex in iRBD (lh)<br>larger volume of fusiform gyrus in iRBD (rh)<br>reduced volume of inferior parietal cortex in iRBD (rh) |
| Deformation | 1 | 0.816<br>(0.140) | 0.714<br>(0.113) | 0.758<br>(0.112) | 0.711<br>(0.132) | Linear SVM, C = 1 | deformation of caudal middle frontal gyrus in iRBD (lh) |
|  | 5 | 0.768<br>(0.149) | 0.852<br>(0.164) | 0.803<br>(0.142) | 0.791<br>(0.152) | Linear SVM, C = 1 | deformation of caudal middle frontal gyrus in iRBD (lh)<br>deformation of paracentral lobule in iRBD (lh)<br>deformation of thalamus proper in iRBD (lh)<br>deformation of putamen in iRBD (lh)<br>deformation of putamen in iRBD (rh) |
| All modalities | 1 | 0.816<br>(0.140) | 0.714<br>(0.113) | 0.758<br>(0.112) | 0.711<br>(0.132) | Linear SVM, C = 1 | deformation of caudal middle frontal gyrus in iRBD (lh) |
|  | 5 | 0.776<br>(0.176) | 0.855<br>(0.170) | 0.806<br>(0.154) | 0.796<br>(0.163) | Linear SVM, C = 1 | deformation of caudal middle frontal gyrus in iRBD (lh)<br>thinning of superior frontal gyrus in iRBD (lh)<br>reduced volume of inferior parietal cortex in iRBD (rh)<br>reduced volume of insula in iRBD (rh)<br>deformation of putamen in iRBD (rh) |

|  | Number of features | Recall | Precision | F1-score | Accuracy | Machine learning model | Brain region and characteristics in iRBD vs PD |
| --- | --- | --- | --- | --- | --- | --- | --- |
| All modalities | 1 | 0.848<br>(0.145) | 0.724<br>(0.073) | 0.776<br>(0.086) | 0.700<br>(0.102) | SVM with RBF kernel, C = 1 | smaller volume of insula in iRBD (rh) |
| All modalities | 2 | 0.880<br>(0.129) | 0.786<br>(0.088) | 0.828<br>(0.099) | 0.775<br>(0.125) | SVM with RBF kernel, C = 1 | smaller surface area of insula in iRBD (rh)<br>lower thinning of entorhinal cortex in iRBD (rh) |
| All modalities | 3 | 0.896<br>(0.131) | 0.801<br>(0.114) | 0.842<br>(0.105) | 0.790<br>(0.134) | SVM with RBF kernel, C = 1 | smaller surface area of insula in iRBD (rh)<br>lower thinning of entorhinal cortex in iRBD (rh)<br>lower thinning of lingual gyrus in iRBD (rh) |
| All modalities | 5 | 0.904<br>(0.131) | 0.832<br>(0.120) | 0.863<br>(0.111) | 0.820<br>(0.140) | SVM with RBF kernel, C = 1 | smaller volume of insula in iRBD (rh)<br>smaller surface area of insula in iRBD (rh)<br>lower thinning of entorhinal cortex in iRBD (rh)<br>lower thinning of lingual gyrus in iRBD (rh)<br>greater volume of fusiform gyrus in iRBD (rh) |
|  | Number of features | Recall | Precision | F1-score | Accuracy | Machine learning model | Brain region and characteristics in iRBD with MCI vs without MCI |
| All modalities | 1 | 0.907<br>(0.205) | 0.525<br>(0.094) | 0.660<br>(0.127) | 0.728<br>(0.079) | Decision tree, depth = 1 | greater thinning of pars triangularis in iRBD with MCI (lh) |
| All modalities | 2 | 0.907<br>(0.181) | 0.537<br>(0.092) | 0.667<br>(0.104) | 0.732<br>(0.075) | Decision tree, depth = 1 | greater thinning of pars triangularis in iRBD with MCI (lh)<br>greater thinning of superior temporal gyrus in iRBD with MCI (lh) |
| All modalities | 3 | 0.733<br>(0.236) | 0.701<br>(0.188) | 0.692<br>(0.160) | 0.812<br>(0.093) | Linear SVM, C = 1 | greater thinning of pars triangularis in iRBD with MCI (lh)<br>greater thinning of transverse temporal cortex in iRBD with MCI (lh)<br>deformation of isthmus of cingulate gyrus in iRBD with MCI (rh) |
| All modalities | 5 | 0.787<br>(0.190) | 0.749<br>(0.171) | 0.754<br>(0.147) | 0.848<br>(0.087) | Linear SVM, C = 1 | greater thinning of pars triangularis in iRBD with MCI (lh)<br>greater thinning of superior temporal gyrus in iRBD with MCI (lh)<br>larger surface area of superior temporal gyrus in iRBD with MCI (lh)<br>greater thinning of transverse temporal cortex in iRBD with MCI (lh)<br>deformation of isthmus of cingulate gyrus in iRBD with MCI (rh) |

Data are shown as mean (SD). All values reported are the average of model performance obtained from the test sets of the 25 randomly generated, unique train-test splits. iRBD: idiopathic rapid eye movement sleep behavior disorder, lh: left hemisphere, MCI: mild cognitive impairment, PD: Parkinson's disease, RBF: radial basis function, SVM: support vector machine, rh: right hemisphere.

Depending on the modality used, linear SVM or SVM with a radial basis function (RBF) was the best performing model in most, if not all, of the 25 randomly generated train-test splits (Table 2; n=24.95 (0.22); range=24-25). As indicated by the relative importance, cortical regions, including the left caudal middle frontal cortex (reduced volume, deformation), paracentral lobule (reduced volume, deformation), and superior frontal cortex (thinning), as well as the right inferior parietal cortex (reduced volume), and insula (reduced volume), yielded high discriminative power for the detection of iRBD (Table 2; Supplementary Tables 2 and 3).

#### iRBD vs PD

With morphometric measurements of 5 brain regions, including, in iRBD, a smaller volume and surface area of the right insula, lower thinning of the right entorhinal cortex and lingual gyrus, and greater volume of the right fusiform gyrus, an accuracy of 82% was achieved (Table 2; Figures 1f and 2). When morphometric measurements of single brain regions were used to train the machine learning model, smaller volume of the right insula in iRBD led to an accuracy of 70% (Table 2). SVM with an RBF kernel demonstrated the highest performance in a majority of the 25 randomly generated train-test splits (n=24.75 (0.5); range=24-25).

#### iRBD with MCI vs iRBD without MCI

An accuracy of 84.8% was obtained from a model trained with all morphometric measurements of 5 brain regions, namely thinning of the left transverse temporal cortex, pars triangularis and superior temporal gyrus, larger surface area of the left superior temporal gyrus, and deformation of isthmus of the right cingulate gyrus in iRBD patients with MCI (Table 2; Figures 1g and 2). When using single brain regions (thinning of the left pars triangularis in iRBD with MCI) or 2 regions (thinning of the left pars triangularis and superior temporal gyrus in iRBD with MCI), a decision tree classifier of depth 1 achieved an accuracy of 72.8% and 73.2%, respectively (Table 2; Figures 1g and 2). When more brain regions were used, a linear SVM yielded the highest overall performance in most of the train-test splits (n=22.5 (0.71); range=22-23).

## Discussion

### General observations

In this study, we derived a comprehensive set of morphological regional brain measurements and used machine learning to identify regions that best discriminate between iRBD patients and controls, iRBD and PD patients, and cognitive subtypes of iRBD. We showed that machine learning models trained with morphometric measurements efficiently differentiate between iRBD and controls (accuracy=79.6%), iRBD and PD (accuracy=82%), and iRBD with or without MCI (accuracy=84.8%) and may prove useful for guiding future algorithms that discriminate patients based on structural MRI.

When discriminating iRBD patients from controls with all morphometric modalities, an accuracy of 79.6% was achieved by models trained using deformation of the left caudal middle frontal gyrus and right putamen, thinning of the left superior frontal gyrus, and reduced volume of the right inferior parietal cortex and insula. With single modalities, an accuracy of 79.1% was obtained from models trained with DBM-derived data, i.e., tissue deformation in the left caudal middle frontal gyrus, paracentral lobule, thalamus, and bilateral putamen. For the other modalities, namely cortical surface-derived measures of thickness, surface area or volume, a lower but above-chance-level accuracy was also observed. This supports the importance of tissue deformation in the structural pattern of atrophy seen in iRBD, as well as the importance of taking into consideration the changes occurring in subcortical regions in iRBD (6,9,10,12–14). The model trained with greater volume and surface area of the right insula, thinning of the right entorhinal cortex and lingual gyrus, and reduced volume of the right fusiform gyrus led to an accuracy of 82% in the discrimination of iRBD from PD patients. Furthermore, an accuracy of 84.8% was obtained in the identification of iRBD patients with MCI, from a model trained with thinning of the left transverse temporal cortex, pars triangularis and superior temporal gyrus, larger surface area of the left superior temporal gyrus, and deformation of isthmus of the right cingulate gyrus.

Few studies have used machine learning to identify iRBD patients with MRI, EEG/PSG, or clinical markers. One study used machine learning models with diffusion tensor imaging measures and found structural differences between 20 iRBD patients and 20 controls, enabling identification of iRBD with an accuracy of 87.5% (26). One study derived features from EEG signals during sleep to train a random forest model that identified iRBD vs controls with an accuracy of 96%, a sensitivity of 98%, and a specificity of 94% (24). In another study that used PSG data, electrooculogram signals were acquired for deriving features relevant to micro-sleep structure, which gave rise to identification of iRBD patients in PD patients through micro-sleep instability, with accuracy, sensitivity, and specificity over 80% (22). In a longitudinal study (23), resting EEG data were analyzed using a convolutional neural network and a recurrent neural network. The two achieved comparable results, leading to an accuracy of 80% when predicting conversion to PD in iRBD. One study used sensor data recorded during motor tasks and trained random forest classifiers to distinguish controls, iRBD and PD patients, and obtained a sensitivity ranging from 84.6% to 91.9%, and a specificity ranging from 88.3% to 90.1% (25). Another study used olfaction data (21) and reported an area under the curve value of ≥0.90 when classifying between iRBD and PD. While these studies demonstrated a high accuracy in the identification of iRBD or PD, many are based on motor features that appear at a later stage of neurodegeneration as in patients at risk of DLB. Moreover, in motor tasks and EEG/PSG, data acquisition can take up to days, making quality control and follow-ups challenging. In addition, previous studies have not attempted to subtype between iRBD patients with or without MCI, for which we have achieved an accuracy of 84.8%. This is essential given the large heterogeneity of cognitive impairment reported in iRBD patients, and the higher risk of developing DLB or PD with cognitive impairment in iRBD patients with MCI. Comparing with these studies, the present study also highlights structural brain changes that allow the identification and subtyping of iRBD.

Deformation of putamen and thalamus, and thinning of caudal middle frontal and paracentral cortices best distinguished between iRBD patients and controls. This is in line with studies performed on the same cohorts using single morphometric measures (12,15,16). Putamen dopamine depletion underlies parkinsonism (45), while the putamen is structurally and functionally abnormal in iRBD (12,46–50), as observed by reduced dopamine reuptake imaging (5,47) and perfusion/metabolism imaging (48–50). The smaller volume of the insula in iRBD was also found to discriminate between iRBD and PD. In the present study, greater thinning of the left insula was observed in iRBD patients with MCI, and the presence of MCI in iRBD is a risk factor for DLB (5,6,8), which is in agreement with studies reporting the insula as highly associated with prodromal and clinical DLB (51–54). Its ability to discriminate between iRBD and PD may therefore be due to some iRBD patients being on a dementia-first trajectory. Also, in accordance with studies that used morphometric measures of the fusiform gyrus to detect PD (55,56), our study highlighted the volume of the fusiform gyrus as a discriminative feature.

Compared to surface-derived cortical metrics assessing local thickness, surface area or volume, DBM-derived tissue deformation measurements generated higher discriminative power between iRBD patients and controls. This is important, as different morphological metrics were shown to follow different developmental trajectories (57), to relate to different genetic determinants (58,59), and to be affected differentially in neurodegenerative prodromes (15,60). Several reasons can explain the fact that DBM is most discriminative. First, DBM maps included information about subcortical structures, which are known to be affected in iRBD (9,10). Secondly, white matter changes are an increasingly recognized feature of neurodegeneration in PD and DLB (61,62); although tissue deformation measurements were extracted from the gray matter only, it is possible that gray matter deformation may have been influenced by dynamics occurring in the underlying white matter. This is even more important as some studies have reported white matter abnormalities in iRBD (14,63,64). Recently, we identified in the same cohort of iRBD patients a DBM-derived structural signature that predicts the development of DLB in iRBD; interestingly, white matter tissue contraction was an important feature of this signature (16). Future studies should investigate the association between changes in gray and white matter in iRBD.

We also found that some of the discriminative brain regions in the classification between iRBD patients and controls, and the identification of PD patients (e.g., the medial orbitofrontal cortex and pars orbitalis) are involved in olfactory processing. Morphometric measures of olfactory processing areas in the orbitofrontal cortex such as the olfactory sulcus, the gyrus rectus, and the medial orbitofrontal cortex are associated with olfactory sensitivity (65,66). Further, olfactory function is impaired in both PD and iRBD patients (3,67). We have shown in a previous study that a convolutional neural network can distinguish PD patients from patients with non-parkinsonian olfactory dysfunction (68). In the future, olfaction and other markers of motor or cognitive performance could be combined with morphometric measures to increase the specificity and sensitivity of iRBD identification (5).

### Limitations

From a clinical perspective, brain imaging data used in this study were collected from a considerable number of participants. However, they represent a relatively small sample size when compared with deep learning studies using multicentric data, or with studies applying machine learning and deep learning to iRBD detection using PSG or EEG features. To enable computer-aided decision-making in the identification and subtyping of iRBD, studies using a larger prospective-longitudinal cohort are required. In addition, it has been shown that the version of the preprocessing pipeline may become a confounding factor in the assessment of neurological disorders (69), therefore, it would be crucial to understand to what extent the detection of iRBD is affected by the preprocessing pipeline, and/or the software version used.

## Data Availability

Data sharing is not applicable to this article as no new data were created or analyzed in this study.

## Author roles

J. Mei: Conception and execution of research project, design and execution of statistical analysis, writing the first draft of manuscript.

SR: Conception of research project, design, review and critique of statistical analysis, writing the first draft of manuscript.

CD: Review and critique of statistical analysis, review of manuscript.

RBP: Funding, organization of research project, data collection, review of manuscript.

J. Montplaisir: Funding, organization of research project, data collection, review of manuscript.

JC: Organization of research project, data collection, review of manuscript.

OM: Organization of research project, data collection, review of manuscript.

JF: Funding, organization and supervision of research project, review of manuscript.

JFG: Funding, organization and supervision of research project, data collection, writing and review of manuscript.

## Acknowledgements

We would like to thank Amélie Pelletier and Jessie De Roy for their assistance in organizing data, and Sylvain Chouinard and Michel Panisset for clinical evaluations of PD patients. Shady Rahayel receives a scholarship from the Fonds de recherche du Québec – Santé. JFG holds a Canada Research Chair in Cognitive Decline in Pathological Aging.

## Financial Disclosures

This work was supported by the Canadian Institutes of Health Research (CIHR), the *Fonds de recherche du Québec – Santé* (FRQ-S), the *Réseau intersectoriel de recherche en santé de l’Université du Québec* (RISUQ), the Centre de recherche en neurosciences cognitives de l’Université du Québec à Montréal (NeuroQAM), and the W. Garfield Weston Foundation. JFG received research grant outside this work from the National Institutes of Health/National Institute for Aging. This work was supported by grants from the Canadian Institutes of Health Research (CIHR, PJT173514) and Parkinson Canada (PPG-2020-0000000061) to JF.

J. Mei:

Stock Ownership in medically-related fields: Oyama Health AB

Consultancies: deepkapha.ai, Oyama Health AB

Employment: Université du Québec à Trois-Rivières, University of Western Ontario

Grants: BrainsCAN Postdoctoral Fellowship Award through Canada First Research Excellence Fund (CFREF)

Partnerships: Charité Global Health

JF:

Employment: Université du Québec à Trois-Rivières

Royalties: Styriabooks

Grants: FRQS, NSERC, CIHR

## Supplementary materials

**Supplementary Table 1.** Morphometric measurements that differed significantly between controls and iRBD patients.

| Morphometric modality and hemisphere | Brain region | Controls | iRBD | P value |
| --- | --- | --- | --- | --- |
| Thickness, left | Superior frontal gyrus | 2.55 (0.08) | 2.48 (0.11) | 0.001 |
|  | Paracentral lobule | 2.39 (0.1) | 2.32 (0.13) | 0.005 |
|  | Temporal pole | 3.53 (0.25) | 3.38 (0.22) | 0.015 |
|  | Inferior temporal gyrus | 2.65 (0.12) | 2.59 (0.12) | 0.017 |
|  | Lateral orbital frontal cortex | 2.52 (0.08) | 2.47 (0.11) | 0.025 |
|  | Middle temporal gyrus | 2.70 (0.1) | 2.65 (0.11) | 0.033 |
|  | Rostral middle frontal gyrus | 2.27 (0.07) | 2.23 (0.08) | 0.034 |
|  | Caudal middle frontal gyrus | 2.45 (0.09) | 2.41 (0.1) | 0.038 |
|  | Pars orbitalis | 2.51 (0.14) | 2.44 (0.16) | 0.038 |
|  | Precentral gyrus | 2.52 (0.09) | 2.46 (0.13) | 0.049 |
| Thickness, right | Superior frontal gyrus | 2.51 (0.08) | 2.46 (0.10) | 0.008 |
|  | Precentral gyrus | 2.50 (0.10) | 2.42 (0.13) | 0.011 |
|  | Supramarginal gyrus | 2.45 (0.08) | 2.40 (0.11) | 0.013 |
|  | Frontal pole | 2.49 (0.18) | 2.41 (0.19) | 0.027 |
|  | Middle temporal gyrus | 2.76 (0.11) | 2.70 (0.11) | 0.031 |
|  | Insula | 2.83 (0.14) | 2.77 (0.13) | 0.034 |
|  | Inferior parietal cortex | 2.39 (0.08) | 2.35 (0.11) | 0.045 |
|  | Inferior temporal gyrus | 2.69 (0.10) | 2.64 (0.11) | 0.045 |
| Surface area, left | Caudal middle frontal gyrus | 0.0016 (0.0002) | 0.0015 (0.0002) | 0.033 |
| Volume, left | Paracentral lobule | 0.0024 (0.0003) | 0.0023 (0.0003) | 0.003 |
|  | Caudal middle frontal gyrus | 0.0042 (0.0007) | 0.0039 (0.0005) | 0.007 |
|  | Precuneus cortex | 0.0064 (0.0006) | 0.0062 (0.0007) | 0.018 |
|  | Rostral anterior cingulate cortex | 0.0018 (0.0003) | 0.0016 (0.0003) | 0.025 |
|  | Rostral middle frontal gyrus | 0.0099 (0.0011) | 0.0094 (0.0010) | 0.025 |
|  | Middle temporal gyrus | 0.0072 (0.0010) | 0.0068 (0.0008) | 0.042 |
| Volume, right | Middle temporal gyrus | 0.0077 (0.0009) | 0.0073 (0.0007) | 0.008 |
|  | Inferior parietal cortex | 0.0094 (0.0010) | 0.0090 (0.0011) | 0.028 |
|  | Superior frontal gyrus | 0.0138 (0.0014) | 0.0134 (0.0012) | 0.036 |
|  | Insula | 0.0049 (0.0006) | 0.0047 (0.0003) | 0.042 |
| Deformation, left | Putamen | 0.94 (0.05) | 0.91 (0.05) | 0.003 |
|  | Caudal middle frontal gyrus | 1.05 (0.08) | 1.01 (0.05) | 0.008 |
|  | Pallidum | 0.94 (0.05) | 0.91 (0.05) | 0.017 |
|  | Rostral middle frontal gyrus | 1.04 (0.04) | 1.02 (0.03) | 0.045 |
|  | Middle temporal gyrus | 1.02 (0.04) | 1.00 (0.04) | 0.048 |
| Deformation, right | Putamen | 0.94 (0.05) | 0.91 (0.05) | 0.001 |
|  | Banks of superior temporal sulcus | 1.02 (0.07) | 0.98 (0.08) | 0.012 |
|  | Pallidum | 0.94 (0.05) | 0.91 (0.05) | 0.035 |

iRBD: idiopathic rapid eye movement sleep behavior disorder. Surface area and volume values were normalized using the estimated total intracranial volume. Red: Thinning, reduced surface area or volume, or tissue deformation value.

**Supplementary Table 2.**
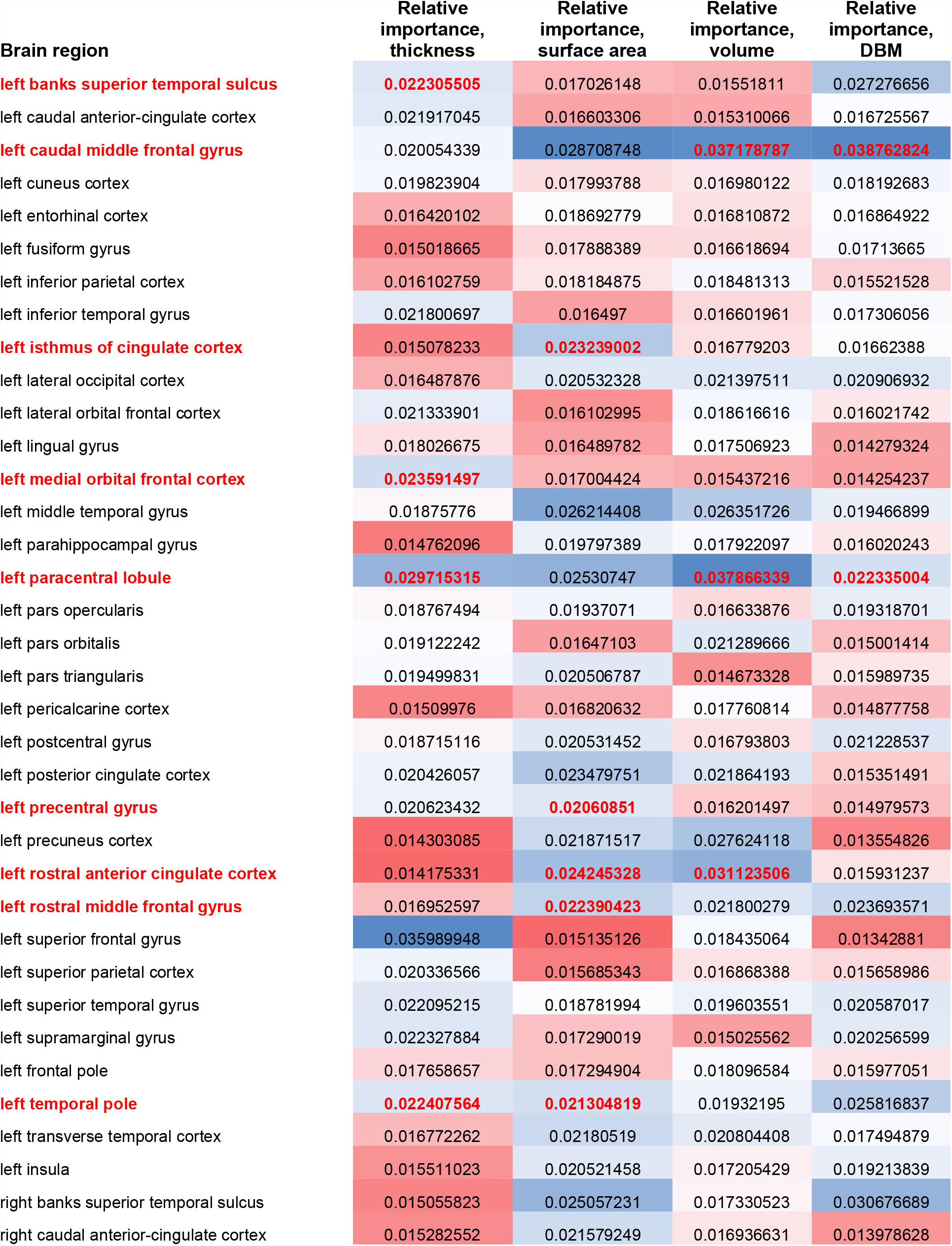
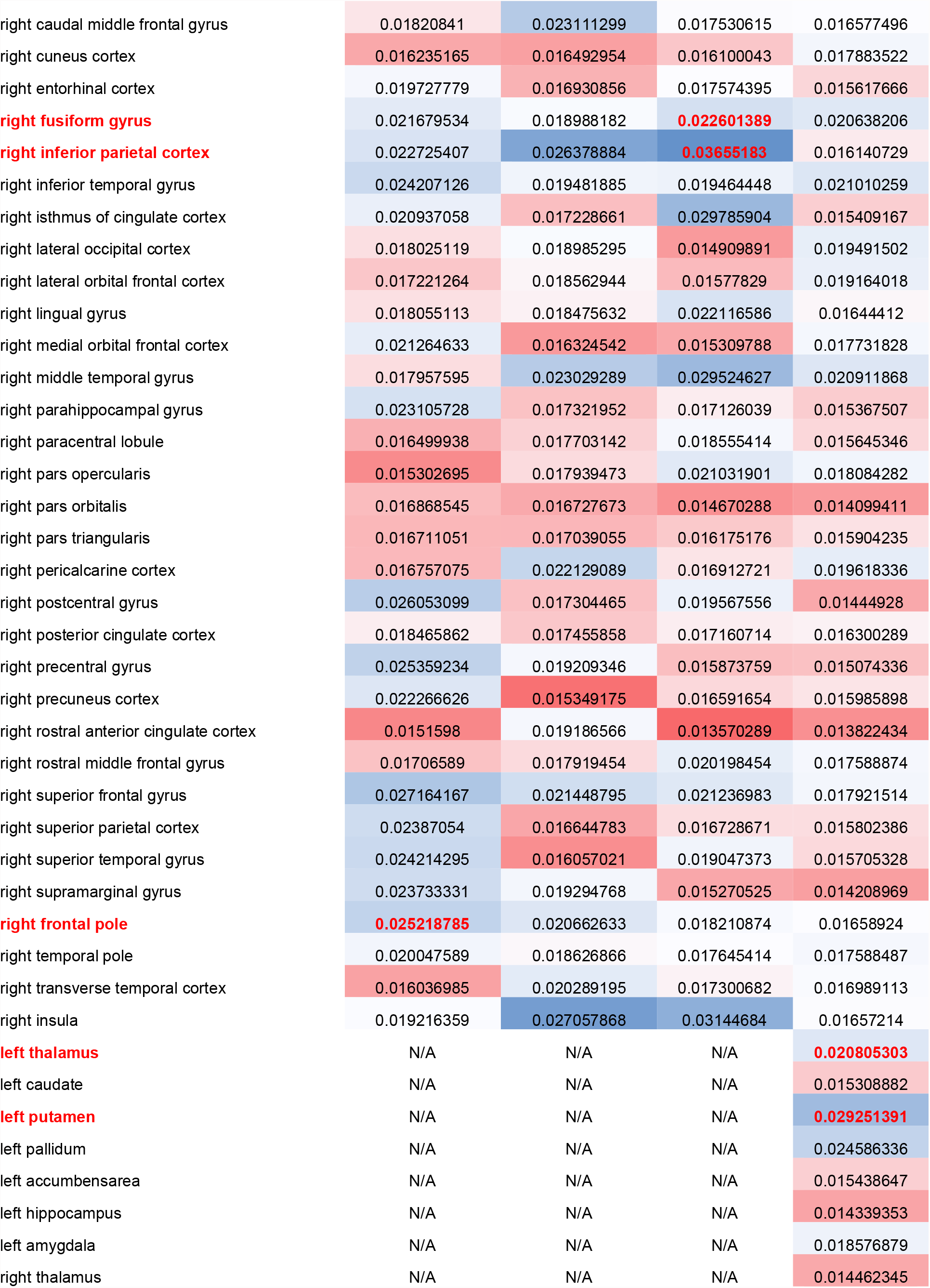
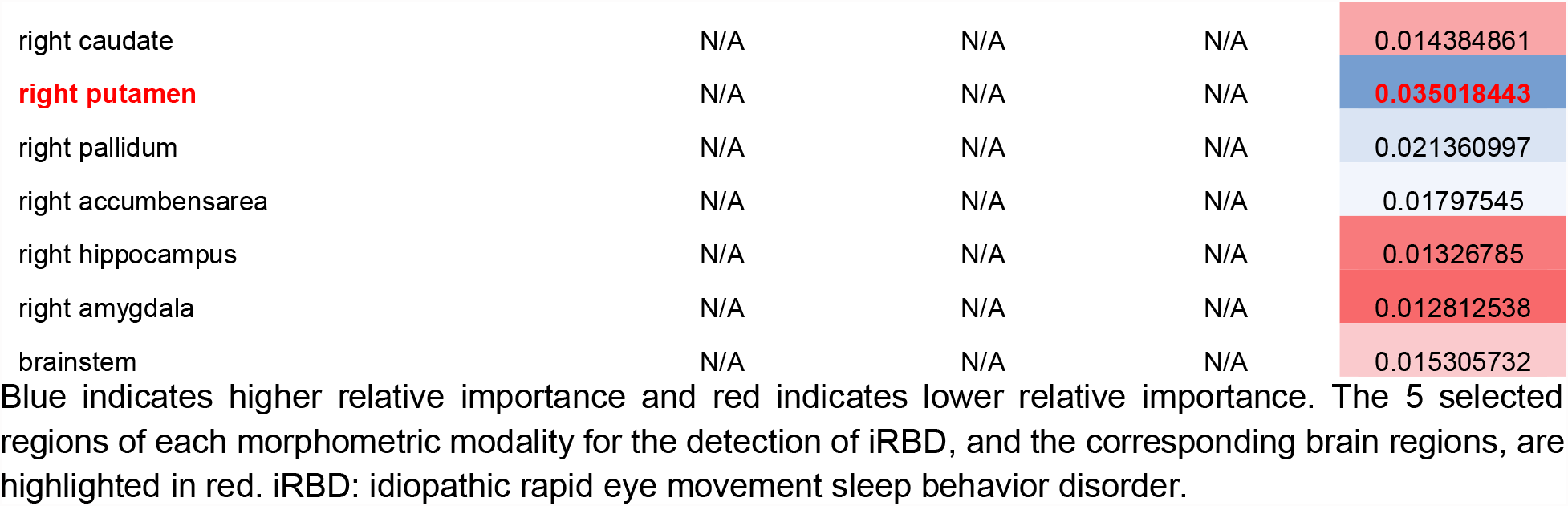
Morphometric measurements of individual modalities and their relative importance in the discrimination between iRBD patients and controls.

**Supplementary Table 3.** Morphometric measurements of all modalities and their relative importance in the three classification tasks.

| Modality | Brain region | Relative importance in iRBD vs controls | Relative importance in iRBD vs PD | Relative importance in iRBD with MCI vs without MCI |
| --- | --- | --- | --- | --- |
| Thickness | left banks superior temporal sulcus | 0.014079179 | 0.019549711 | 0.039856179 |
| Thickness | left caudal anterior-cingulate cortex | 0.013323188 | 0.017439446 | 0.030950973 |
| Thickness | left caudal middle frontal gyrus | 0.015908295 | 0.014067108 | 0.032876387 |
| Thickness | left cuneus cortex | 0.012438472 | 0.020779845 | 0.027326406 |
| Thickness | left entorhinal cortex | 0.011722899 | 0.020844082 | 0.030215824 |
| Thickness | left fusiform gyrus | 0.011117685 | 0.021353284 | 0.040265091 |
| Thickness | left inferior parietal cortex | 0.011371211 | 0.019143772 | 0.027155903 |
| Thickness | left inferior temporal gyrus | 0.017070415 | 0.014363645 | 0.031567695 |
| Thickness | left isthmus of cingulate cortex | 0.011348432 | 0.014659224 | 0.036294859 |
| Thickness | left lateral occipital cortex | 0.012300894 | 0.013603443 | 0.027268546 |
| Thickness | left lateral orbital frontal cortex | 0.016928141 | 0.019995745 | 0.030193138 |
| Thickness | left lingual gyrus | 0.011940763 | 0.024650088 | 0.023914679 |
| Thickness | left medial orbital frontal cortex | 0.017097566 | 0.015780046 | 0.022602326 |
| Thickness | left middle temporal gyrus | 0.015410157 | 0.014894575 | 0.052712822 |
| Thickness | left parahippocampal gyrus | 0.011322868 | 0.013668491 | 0.026288787 |
| Thickness | left paracentral lobule | 0.020753132 | 0.013227169 | 0.037548988 |
| Thickness | left pars opercularis | 0.015387613 | 0.018745183 | 0.031698277 |
| Thickness | left pars orbitalis | 0.015690116 | 0.021436755 | 0.039461592 |
| Thickness | **left pars triangularis** | 0.015793965 | 0.013653322 | **0.056895199** |
| Thickness | left pericalcarine cortex | 0.010990394 | 0.014033361 | 0.021892541 |
| Thickness | left postcentral gyrus | 0.012979236 | 0.015893163 | 0.036431948 |
| Thickness | left posterior cingulate cortex | 0.013707549 | 0.017613172 | 0.022848548 |
| Thickness | left precentral gyrus | 0.015142655 | 0.012640453 | 0.031494912 |
| Thickness | left precuneus cortex | 0.011822649 | 0.013012484 | 0.030116075 |
| Thickness | left rostral anterior cingulate cortex | 0.011416324 | 0.016209826 | 0.033126907 |
| Thickness | left rostral middle frontal gyrus | 0.014749473 | 0.034015919 | 0.024428467 |
| Thickness | **left superior frontal gyrus** | **0.025514954** | 0.022849732 | 0.035726972 |
| Thickness | left superior parietal cortex | 0.014691874 | 0.013201828 | 0.03084317 |
| Thickness | **left superior temporal gyrus** | 0.017493471 | 0.018006544 | **0.043440949** |
| Thickness | left supramarginal gyrus | 0.015530589 | 0.015291581 | 0.030474569 |
| Thickness | left frontal pole | 0.012287079 | 0.016768059 | 0.032179412 |
| Thickness | left temporal pole | 0.017097452 | 0.021108641 | 0.025326965 |
| Thickness | **left transverse temporal cortex** | 0.011206907 | 0.016753939 | **0.044647084** |
| Thickness | left insula | 0.012066725 | 0.014303357 | 0.043471837 |
| Thickness | right banks superior temporal sulcus | 0.011735428 | 0.014717026 | 0.033580899 |
| Thickness | right caudal anterior-cingulate cortex | 0.012268814 | 0.017172073 | 0.026779746 |
| Thickness | right caudal middle frontal gyrus | 0.014002181 | 0.013954758 | 0.027208521 |
| Thickness | right cuneus cortex | 0.012509784 | 0.014249984 | 0.035848189 |
| Thickness | **right entorhinal cortex** | 0.013100886 | **0.023520966** | 0.028327844 |
| Thickness | right fusiform gyrus | 0.013564742 | 0.022477871 | 0.023139147 |
| Thickness | right inferior parietal cortex | 0.017054597 | 0.014196839 | 0.024699917 |
| Thickness | right inferior temporal gyrus | 0.018581649 | 0.015156914 | 0.04246015 |
| Thickness | right isthmus of cingulate cortex | 0.016132018 | 0.016527631 | 0.024565563 |
| Thickness | right lateral occipital cortex | 0.012407566 | 0.013661633 | 0.034691799 |
| Thickness | right lateral orbital frontal cortex | 0.011851284 | 0.02041409 | 0.022329636 |
| Thickness | **right lingual gyrus** | 0.014716984 | **0.026008193** | 0.023324283 |
| Thickness | right medial orbital frontal cortex | 0.015246822 | 0.032514829 | 0.021846683 |
| Thickness | right middle temporal gyrus | 0.014854105 | 0.018723 | 0.044885561 |
| Thickness | right parahippocampal gyrus | 0.017089722 | 0.014476989 | 0.027128665 |
| Thickness | right paracentral lobule | 0.012402912 | 0.014473022 | 0.042459882 |
| Thickness | right pars opercularis | 0.01337562 | 0.016224865 | 0.021944947 |
| Thickness | right pars orbitalis | 0.012329492 | 0.017064422 | 0.032894184 |
| Thickness | right pars triangularis | 0.01167223 | 0.023241623 | 0.040046996 |
| Thickness | right pericalcarine cortex | 0.01253267 | 0.017935254 | 0.028649658 |
| Thickness | right postcentral gyrus | 0.016905022 | 0.012458152 | 0.039057056 |
| Thickness | right posterior cingulate cortex | 0.014214478 | 0.023800418 | 0.023898822 |
| Thickness | right precentral gyrus | 0.01877325 | 0.013969073 | 0.033584866 |
| Thickness | right precuneus cortex | 0.016414814 | 0.013399446 | 0.02763449 |
| Thickness | right rostral anterior cingulate cortex | 0.011465599 | 0.013708028 | 0.031977432 |
| Thickness | right rostral middle frontal gyrus | 0.013567612 | 0.021111272 | 0.024116962 |
| Thickness | right superior frontal gyrus | 0.021262416 | 0.016994278 | 0.024228412 |
| Thickness | right superior parietal cortex | 0.016248348 | 0.014227648 | 0.030410239 |
| Thickness | right superior temporal gyrus | 0.01645094 | 0.015434882 | 0.034222454 |
| Thickness | right supramarginal gyrus | 0.018551794 | 0.017093096 | 0.02892838 |
| Thickness | right frontal pole | 0.018494782 | 0.024654214 | 0.035391347 |
| Thickness | right temporal pole | 0.014120754 | 0.01574443 | 0.028458481 |
| Thickness | right transverse temporal cortex | 0.011560923 | 0.018638003 | 0.038637662 |
| Thickness | right insula | 0.015555332 | 0.022126482 | 0.024140652 |
| Surface area | left banks superior temporal sulcus | 0.011804736 | 0.014179616 | 0.028269495 |
| Surface area | left caudal anterior-cingulate cortex | 0.01185018 | 0.01859055 | 0.029566776 |
| Surface area | left caudal middle frontal gyrus | 0.018512643 | 0.013035388 | 0.03350493 |
| Surface area | left cuneus cortex | 0.012769615 | 0.013405796 | 0.027502303 |
| Surface area | left entorhinal cortex | 0.012240109 | 0.013116294 | 0.023918715 |
| Surface area | left fusiform gyrus | 0.011909125 | 0.02103983 | 0.035551085 |
| Surface area | left inferior parietal cortex | 0.013189596 | 0.013833195 | 0.023196357 |
| Surface area | left inferior temporal gyrus | 0.011866603 | 0.016749538 | 0.025251852 |
| Surface area | left isthmus of cingulate cortex | 0.015958487 | 0.0212881 | 0.025904815 |
| Surface area | left lateral occipital cortex | 0.013954291 | 0.014115729 | 0.020282852 |
| Surface area | left lateral orbital frontal cortex | 0.011776968 | 0.014264613 | 0.026393873 |
| Surface area | left lingual gyrus | 0.011346989 | 0.016227995 | 0.025677163 |
| Surface area | left medial orbital frontal cortex | 0.012088102 | 0.012660718 | 0.022317944 |
| Surface area | left middle temporal gyrus | 0.016399823 | 0.015984513 | 0.023282792 |
| Surface area | left parahippocampal gyrus | 0.013368721 | 0.01504468 | 0.026338249 |
| Surface area | left paracentral lobule | 0.01699631 | 0.019962389 | 0.033569856 |
| Surface area | left pars opercularis | 0.01306759 | 0.015761246 | 0.025735661 |
| Surface area | left pars orbitalis | 0.011664572 | 0.013192873 | 0.029988303 |
| Surface area | left pars triangularis | 0.014325188 | 0.019404746 | 0.029884263 |
| Surface area | left pericalcarine cortex | 0.011597313 | 0.013488359 | 0.031033441 |
| Surface area | left postcentral gyrus | 0.013699635 | 0.015344947 | 0.024285691 |
| Surface area | left posterior cingulate cortex | 0.015272823 | 0.013019238 | 0.025061639 |
| Surface area | left precentral gyrus | 0.01457001 | 0.017495095 | 0.026014717 |
| Surface area | left precuneus cortex | 0.014872035 | 0.02078623 | 0.031523674 |
| Surface area | left rostral anterior cingulate cortex | 0.015888193 | 0.014293146 | 0.024590174 |
| Surface area | left rostral middle frontal gyrus | 0.015299064 | 0.015526055 | 0.021310915 |
| Surface area | left superior frontal gyrus | 0.010837738 | 0.016967898 | 0.023230501 |
| Surface area | left superior parietal cortex | 0.011009514 | 0.014980647 | 0.025358578 |
| Surface area | **left superior temporal gyrus** | 0.012902697 | 0.015671332 | **0.0389589** |
| Surface area | left supramarginal gyrus | 0.011926347 | 0.017335015 | 0.020631152 |
| Surface area | left frontal pole | 0.012654333 | 0.015951619 | 0.023122464 |
| Surface area | left temporal pole | 0.01444723 | 0.01549076 | 0.028519145 |
| Surface area | left transverse temporal cortex | 0.014312613 | 0.017301175 | 0.029737349 |
| Surface area | left insula | 0.013577311 | 0.014702295 | 0.031563462 |
| Surface area | right banks superior temporal sulcus | 0.016863066 | 0.014034919 | 0.033839343 |
| Surface area | right caudal anterior-cingulate cortex | 0.015152306 | 0.013328578 | 0.027242533 |
| Surface area | right caudal middle frontal gyrus | 0.013912254 | 0.01455872 | 0.022370967 |
| Surface area | right cuneus cortex | 0.012085357 | 0.01442942 | 0.027223683 |
| Surface area | right entorhinal cortex | 0.011726925 | 0.012861287 | 0.031402504 |
| Surface area | right fusiform gyrus | 0.01278161 | 0.021168499 | 0.033555707 |
| Surface area | right inferior parietal cortex | 0.017478591 | 0.016535118 | 0.029827256 |
| Surface area | right inferior temporal gyrus | 0.014145474 | 0.020408667 | 0.027871193 |
| Surface area | right isthmus of cingulate cortex | 0.011884422 | 0.01784246 | 0.029194251 |
| Surface area | right lateral occipital cortex | 0.01306609 | 0.015053386 | 0.021983859 |
| Surface area | right lateral orbital frontal cortex | 0.013209152 | 0.012999835 | 0.024088024 |
| Surface area | right lingual gyrus | 0.012501777 | 0.014408485 | 0.029635258 |
| Surface area | right medial orbital frontal cortex | 0.011933068 | 0.016890827 | 0.021076271 |
| Surface area | right middle temporal gyrus | 0.015467286 | 0.015510372 | 0.027075188 |
| Surface area | right parahippocampal gyrus | 0.012023279 | 0.017357431 | 0.027573668 |
| Surface area | right paracentral lobule | 0.012697262 | 0.013617372 | 0.0319222 |
| Surface area | right pars opercularis | 0.01212243 | 0.018843115 | 0.023587593 |
| Surface area | right pars orbitalis | 0.011539456 | 0.01516969 | 0.02398069 |
| Surface area | right pars triangularis | 0.012133721 | 0.013781044 | 0.028122856 |
| Surface area | right pericalcarine cortex | 0.013336264 | 0.013580531 | 0.023448461 |
| Surface area | right postcentral gyrus | 0.012220051 | 0.013761146 | 0.022407599 |
| Surface area | right posterior cingulate cortex | 0.01260807 | 0.013966887 | 0.028512021 |
| Surface area | right precentral gyrus | 0.012877169 | 0.015832302 | 0.027681478 |
| Surface area | right precuneus cortex | 0.011143898 | 0.013881345 | 0.035642193 |
| Surface area | right rostral anterior cingulate cortex | 0.013272521 | 0.015343536 | 0.023819673 |
| Surface area | right rostral middle frontal gyrus | 0.012167336 | 0.024348844 | 0.037412131 |
| Surface area | right superior frontal gyrus | 0.014237726 | 0.013254355 | 0.021973606 |
| Surface area | right superior parietal cortex | 0.011395526 | 0.014338586 | 0.037711232 |
| Surface area | right superior temporal gyrus | 0.011665648 | 0.021516198 | 0.023668534 |
| Surface area | right supramarginal gyrus | 0.013394192 | 0.013299528 | 0.027350884 |
| Surface area | right frontal pole | 0.014360818 | 0.017266401 | 0.029843217 |
| Surface area | right temporal pole | 0.01334236 | 0.013353976 | 0.023126946 |
| Surface area | right transverse temporal cortex | 0.01336309 | 0.014229008 | 0.023982466 |
| Surface area | **right insula** | 0.016258779 | **0.025296632** | 0.02487458 |
| Volume | left banks superior temporal sulcus | 0.01163617 | 0.014304678 | 0.023475361 |
| Volume | left caudal anterior-cingulate cortex | 0.010949134 | 0.014700795 | 0.036370093 |
| Volume | left caudal middle frontal gyrus | 0.024401998 | 0.014446482 | 0.031566332 |
| Volume | left cuneus cortex | 0.012598747 | 0.013295854 | 0.023791372 |
| Volume | left entorhinal cortex | 0.011697937 | 0.012764738 | 0.025155303 |
| Volume | left fusiform gyrus | 0.012193735 | 0.02355814 | 0.030813691 |
| Volume | left inferior parietal cortex | 0.013478203 | 0.013416898 | 0.023191798 |
| Volume | left inferior temporal gyrus | 0.012192352 | 0.018309871 | 0.025962536 |
| Volume | left isthmus of cingulate cortex | 0.011947355 | 0.017714636 | 0.025129355 |
| Volume | left lateral occipital cortex | 0.01522789 | 0.015730507 | 0.021918332 |
| Volume | left lateral orbital frontal cortex | 0.014266613 | 0.015229908 | 0.021579694 |
| Volume | left lingual gyrus | 0.01271458 | 0.019659521 | 0.025987493 |
| Volume | left medial orbital frontal cortex | 0.011034274 | 0.014329334 | 0.022748933 |
| Volume | left middle temporal gyrus | 0.017221981 | 0.016602778 | 0.020612203 |
| Volume | left parahippocampal gyrus | 0.012548249 | 0.014730191 | 0.025405655 |
| Volume | left paracentral lobule | 0.024778782 | 0.017121263 | 0.021159391 |
| Volume | left pars opercularis | 0.012187552 | 0.013854126 | 0.023171459 |
| Volume | left pars orbitalis | 0.015518436 | 0.020967369 | 0.020793319 |
| Volume | left pars triangularis | 0.011315228 | 0.018658424 | 0.025683489 |
| Volume | left pericalcarine cortex | 0.012920082 | 0.013049061 | 0.027496995 |
| Volume | left postcentral gyrus | 0.012876342 | 0.014906299 | 0.031058686 |
| Volume | left posterior cingulate cortex | 0.015052976 | 0.01418775 | 0.03070179 |
| Volume | left precentral gyrus | 0.013001932 | 0.01404499 | 0.022697306 |
| Volume | left precuneus cortex | 0.019891012 | 0.015616971 | 0.022750903 |
| Volume | left rostral anterior cingulate cortex | 0.020981126 | 0.018706983 | 0.022336571 |
| Volume | left rostral middle frontal gyrus | 0.015871038 | 0.018796474 | 0.023478204 |
| Volume | left superior frontal gyrus | 0.014434799 | 0.01724332 | 0.022974848 |
| Volume | left superior parietal cortex | 0.011783881 | 0.013500984 | 0.024786328 |
| Volume | left superior temporal gyrus | 0.015408704 | 0.015510599 | 0.026953967 |
| Volume | left supramarginal gyrus | 0.011522122 | 0.020662282 | 0.024785635 |
| Volume | left frontal pole | 0.013374742 | 0.014815862 | 0.027308285 |
| Volume | left temporal pole | 0.014150304 | 0.018647726 | 0.024455756 |
| Volume | left transverse temporal cortex | 0.01374447 | 0.018369561 | 0.028298155 |
| Volume | left insula | 0.01217071 | 0.01472274 | 0.0253114 |
| Volume | right banks superior temporal sulcus | 0.013073473 | 0.017001092 | 0.024171755 |
| Volume | right caudal anterior-cingulate cortex | 0.013246974 | 0.014727398 | 0.023812067 |
| Volume | right caudal middle frontal gyrus | 0.01208595 | 0.013710942 | 0.02330445 |
| Volume | right cuneus cortex | 0.011473418 | 0.013178819 | 0.022201736 |
| Volume | right entorhinal cortex | 0.012413295 | 0.015339423 | 0.037047848 |
| Volume | **right fusiform gyrus** | 0.015864219 | **0.031892986** | 0.036795807 |
| Volume | **right inferior parietal cortex** | **0.024129616** | 0.017402507 | 0.026931811 |
| Volume | right inferior temporal gyrus | 0.014520287 | 0.018309896 | 0.023086281 |
| Volume | right isthmus of cingulate cortex | 0.01921661 | 0.018458416 | 0.028386609 |
| Volume | right lateral occipital cortex | 0.011076955 | 0.018065395 | 0.024462789 |
| Volume | right lateral orbital frontal cortex | 0.012490477 | 0.01674574 | 0.022536365 |
| Volume | right lingual gyrus | 0.015044684 | 0.017413527 | 0.027544467 |
| Volume | right medial orbital frontal cortex | 0.011896461 | 0.012634484 | 0.021296098 |
| Volume | right middle temporal gyrus | 0.020168018 | 0.015632793 | 0.021764322 |
| Volume | right parahippocampal gyrus | 0.012209865 | 0.019874645 | 0.028151478 |
| Volume | right paracentral lobule | 0.013771375 | 0.016760677 | 0.023085602 |
| Volume | right pars opercularis | 0.013960044 | 0.020564297 | 0.028574734 |
| Volume | right pars orbitalis | 0.011454239 | 0.014614335 | 0.022464252 |
| Volume | right pars triangularis | 0.011098119 | 0.013723619 | 0.022957657 |
| Volume | right pericalcarine cortex | 0.012562483 | 0.01596943 | 0.023239643 |
| Volume | right postcentral gyrus | 0.014665665 | 0.013871988 | 0.027270573 |
| Volume | right posterior cingulate cortex | 0.011608123 | 0.012336501 | 0.024783097 |
| Volume | right precentral gyrus | 0.012587535 | 0.013019264 | 0.020992576 |
| Volume | right precuneus cortex | 0.011828327 | 0.015049186 | 0.027861762 |
| Volume | right rostral anterior cingulate cortex | 0.010981158 | 0.015875024 | 0.023987197 |
| Volume | right rostral middle frontal gyrus | 0.014935121 | 0.025707931 | 0.02941208 |
| Volume | right superior frontal gyrus | 0.016122175 | 0.0164729 | 0.024702794 |
| Volume | right superior parietal cortex | 0.012020065 | 0.014489253 | 0.023803039 |
| Volume | right superior temporal gyrus | 0.014277264 | 0.015745437 | 0.02359007 |
| Volume | right supramarginal gyrus | 0.011621179 | 0.012922611 | 0.021889786 |
| Volume | right frontal pole | 0.013687657 | 0.025410771 | 0.02934109 |
| Volume | right temporal pole | 0.012320566 | 0.014939848 | 0.023471438 |
| Volume | right transverse temporal cortex | 0.012489608 | 0.012981911 | 0.024938289 |
| Volume | **right insula** | **0.021323892** | **0.027336652** | 0.025125714 |
| Deformation | left banks superior temporal sulcus | 0.020139406 | 0.014448023 | 0.023410098 |
| Deformation | left caudal anterior-cingulate cortex | 0.012518083 | 0.014971862 | 0.023583531 |
| Deformation | **left caudal middle frontal gyrus** | **0.026359012** | 0.022439221 | 0.02198052 |
| Deformation | left cuneus cortex | 0.015035229 | 0.014231586 | 0.028490458 |
| Deformation | left entorhinal cortex | 0.01323186 | 0.01294874 | 0.027161767 |
| Deformation | left fusiform gyrus | 0.014534996 | 0.014234563 | 0.02199874 |
| Deformation | left inferior parietal cortex | 0.011808512 | 0.013867425 | 0.024679826 |
| Deformation | left inferior temporal gyrus | 0.014119562 | 0.016837995 | 0.02948465 |
| Deformation | left isthmus of cingulate cortex | 0.013517441 | 0.016947317 | 0.030175269 |
| Deformation | left lateral occipital cortex | 0.015955581 | 0.019697677 | 0.02135947 |
| Deformation | left lateral orbital frontal cortex | 0.012147063 | 0.016011764 | 0.022333903 |
| Deformation | left lingual gyrus | 0.011536185 | 0.017011561 | 0.022233982 |
| Deformation | left medial orbital frontal cortex | 0.011217434 | 0.024784672 | 0.025018164 |
| Deformation | left middle temporal gyrus | 0.016187467 | 0.014148576 | 0.028300535 |
| Deformation | left parahippocampal gyrus | 0.012369806 | 0.017078687 | 0.037010084 |
| Deformation | left paracentral lobule | 0.016176805 | 0.021102808 | 0.031619591 |
| Deformation | left pars opercularis | 0.014941853 | 0.013315829 | 0.022823418 |
| Deformation | left pars orbitalis | 0.011568726 | 0.015501357 | 0.029512368 |
| Deformation | left pars triangularis | 0.012192376 | 0.017883305 | 0.026281511 |
| Deformation | left pericalcarine cortex | 0.012191062 | 0.013144696 | 0.021064911 |
| Deformation | left postcentral gyrus | 0.017498254 | 0.012547789 | 0.026571902 |
| Deformation | left posterior cingulate cortex | 0.012396343 | 0.032003898 | 0.031165804 |
| Deformation | left precentral gyrus | 0.011497233 | 0.013397404 | 0.025165174 |
| Deformation | left precuneus cortex | 0.011212541 | 0.017388392 | 0.022359431 |
| Deformation | left rostral anterior cingulate cortex | 0.01279649 | 0.017073704 | 0.022159183 |
| Deformation | left rostral middle frontal gyrus | 0.017125414 | 0.01929352 | 0.035351476 |
| Deformation | left superior frontal gyrus | 0.010788915 | 0.016492326 | 0.028676281 |
| Deformation | left superior parietal cortex | 0.011943207 | 0.014287664 | 0.021282456 |
| Deformation | left superior temporal gyrus | 0.01602699 | 0.016573515 | 0.025785755 |
| Deformation | left supramarginal gyrus | 0.016132979 | 0.013050433 | 0.023144003 |
| Deformation | left frontal pole | 0.012682229 | 0.014324081 | 0.024920648 |
| Deformation | left temporal pole | 0.018384577 | 0.014301561 | 0.023326803 |
| Deformation | left transverse temporal cortex | 0.013562159 | 0.015069344 | 0.026169376 |
| Deformation | left insula | 0.014360485 | 0.016721512 | 0.035604736 |
| Deformation | right banks superior temporal sulcus | 0.02246122 | 0.0132415 | 0.026333088 |
| Deformation | right caudal anterior-cingulate cortex | 0.011695458 | 0.012276017 | 0.022584054 |
| Deformation | right caudal middle frontal gyrus | 0.01259305 | 0.019684074 | 0.024045628 |
| Deformation | right cuneus cortex | 0.014762746 | 0.01353393 | 0.024798605 |
| Deformation | right entorhinal cortex | 0.012181006 | 0.013245076 | 0.026334405 |
| Deformation | right fusiform gyrus | 0.015518617 | 0.014724211 | 0.026507709 |
| Deformation | right inferior parietal cortex | 0.012860464 | 0.018053868 | 0.022338962 |
| Deformation | right inferior temporal gyrus | 0.01733522 | 0.0133652 | 0.022688264 |
| Deformation | right isthmus of cingulate cortex | 0.01241632 | 0.016685857 | 0.039285114 |
| Deformation | right lateral occipital cortex | 0.014731728 | 0.013113821 | 0.027053321 |
| Deformation | right lateral orbital frontal cortex | 0.014701487 | 0.013380676 | 0.024966149 |
| Deformation | right lingual gyrus | 0.013118936 | 0.017885638 | 0.021338641 |
| Deformation | right medial orbital frontal cortex | 0.014793091 | 0.015301727 | 0.022287184 |
| Deformation | right middle temporal gyrus | 0.017034367 | 0.014137359 | 0.025590129 |
| Deformation | right parahippocampal gyrus | 0.011936899 | 0.013551665 | 0.032585538 |
| Deformation | right paracentral lobule | 0.011903223 | 0.012701943 | 0.032846536 |
| Deformation | right pars opercularis | 0.013533817 | 0.013998113 | 0.032443056 |
| Deformation | right pars orbitalis | 0.011231039 | 0.02009125 | 0.028041412 |
| Deformation | right pars triangularis | 0.012333417 | 0.01750689 | 0.023309404 |
| Deformation | right pericalcarine cortex | 0.015503081 | 0.0128513 | 0.023601159 |
| Deformation | right postcentral gyrus | 0.011489251 | 0.014301173 | 0.02341984 |
| Deformation | right posterior cingulate cortex | 0.012926597 | 0.017864468 | 0.023355027 |
| Deformation | right precentral gyrus | 0.011740956 | 0.015612647 | 0.02123393 |
| Deformation | right precuneus cortex | 0.012794881 | 0.016704011 | 0.023945819 |
| Deformation | right rostral anterior cingulate cortex | 0.010942776 | 0.012748099 | 0.023831637 |
| Deformation | right rostral middle frontal gyrus | 0.013931446 | 0.02327875 | 0.03940495 |
| Deformation | right superior frontal gyrus | 0.01376992 | 0.013001065 | 0.024142899 |
| Deformation | right superior parietal cortex | 0.012356985 | 0.020700551 | 0.022328288 |
| Deformation | right superior temporal gyrus | 0.012290483 | 0.015964187 | 0.028270222 |
| Deformation | right supramarginal gyrus | 0.010969915 | 0.013459052 | 0.022989722 |
| Deformation | right frontal pole | 0.012854473 | 0.014403872 | 0.030211319 |
| Deformation | right temporal pole | 0.014006673 | 0.016264848 | 0.022342454 |
| Deformation | right transverse temporal cortex | 0.012921095 | 0.013714227 | 0.030880462 |
| Deformation | right insula | 0.013508908 | 0.015752182 | 0.024881475 |
| Deformation | left thalamus | 0.016088914 | 0.014288133 | 0.025419124 |
| Deformation | left caudate | 0.012198555 | 0.012931672 | 0.025281209 |
| Deformation | left putamen | 0.021505507 | 0.025581131 | 0.033742741 |
| Deformation | left pallidum | 0.018876772 | 0.020081253 | 0.039167822 |
| Deformation | left accumbens area | 0.012475014 | 0.0192723 | 0.025813701 |
| Deformation | left hippocampus | 0.011783754 | 0.014219076 | 0.02916447 |
| Deformation | left amygdala | 0.015219171 | 0.012917429 | 0.030234155 |
| Deformation | right thalamus | 0.011626025 | 0.015914864 | 0.028790682 |
| Deformation | right caudate | 0.011884908 | 0.01183014 | 0.024841387 |
| Deformation | **right putamen** | **0.024620056** | 0.019946589 | 0.034625928 |
| Deformation | right pallidum | 0.016529018 | 0.014116426 | 0.034465437 |
| Deformation | right accumbens area | 0.014147189 | 0.015978305 | 0.027909382 |
| Deformation | right hippocampus | 0.011570919 | 0.013928616 | 0.030238036 |
| Deformation | right amygdala | 0.010849397 | 0.014988168 | 0.027716113 |
| Deformation | brainstem | 0.011710106 | 0.013054873 | 0.023888922 |

Blue indicates higher relative importance and red indicates lower relative importance. The 5 selected regions in each classification, and the corresponding brain regions, are highlighted in red. iRBD: idiopathic rapid eye movement sleep behavior disorder. PD: Parkinson's disease. MCI: mild cognitive impairment.

**Supplementary Table 4.** Morphometric measurements that differed significantly between PD and iRBD patients.

| Morphometric modality and hemisphere | Brain region | PD | iRBD | P value |
| --- | --- | --- | --- | --- |
| Thickness, left | Lingual gyrus | 1.95 (0.08) | 2.00 (0.08) | 0.003 |
|  | Rostral middle frontal gyrus | 2.29 (0.09) | 2.23 (0.08) | 0.005 |
|  | Superior frontal gyrus | 2.55 (0.08) | 2.48 (0.11) | 0.009 |
|  | Pars orbitalis | 2.52 (0.14) | 2.44 (0.16) | 0.046 |
| Thickness, right | Medial orbital frontal cortex | 2.33 (0.14) | 2.24 (0.10) | 0.003 |
|  | Frontal pole | 2.56 (0.25) | 2.41 (0.19) | 0.005 |
|  | Entorhinal cortex | 2.99 (0.23) | 3.13 (0.19) | 0.013 |
|  | Pericalcarine cortex | 1.72 (0.13) | 1.79 (0.12) | 0.028 |
|  | Posterior cingulate cortex | 2.33 (0.15) | 2.26 (0.11) | 0.033 |
|  | Pars triangularis | 2.30 (0.12) | 2.26 (0.11) | 0.040 |
|  | Lingual gyrus | 1.98 (0.10) | 2.02 (0.08) | 0.043 |
| Surface area, left | Isthmus of cingulate cortex | 0.00071 (0.0001) | 0.00075 (0.0001) | 0.046 |
| Volume, left | Fusiform gyrus | 0.0057 (0.0005) | 0.0060 (0.0006) | 0.020 |
|  | Lingual gyrus | 0.0040 (0.0006) | 0.0043 (0.0006) | 0.025 |
| Volume, right | Fusiform gyrus | 0.0057 (0.0005) | 0.0062 (0.0009) | 0.002 |
|  | Frontal pole | 0.0007 (0.0001) | 0.0007 (0.0001) | 0.04992 |
| Deformation, left | Posterior cingulate cortex | 1.00 (0.06) | 0.97 (0.05) | 0.007 |
|  | Pallidum | 0.93 (0.06) | 0.91 (0.05) | 0.039 |
|  | Accumbens area | 1.01 (0.04) | 0.99 (0.05) | 0.049 |
|  | Putamen | 0.93 (0.06) | 0.91 (0.05) | 0.04992 |

PD: Parkinson's disease, iRBD: idiopathic rapid eye movement sleep behavior disorder. Surface area and volume values were normalized using the estimated total intracranial volume. Red: Thinning, reduced surface area or volume, or tissue deformation value.

**Supplementary Table 5.** Morphometric measurements that differed significantly between iRBD patients with or without MCI.

| Morphometric modality and hemisphere | Brain region | iRBD without MCI | iRBD with MCI | P value |
| --- | --- | --- | --- | --- |
| Thickness, left | Pars triangularis | 2.33 (0.11) | 2.21 (0.06) | 0.0002 |
|  | Pars orbitalis | 2.49 (0.16) | 2.35 (0.12) | 0.005 |
|  | Superior temporal gyrus | 2.64 (0.11) | 2.53 (0.14) | 0.006 |
|  | Transverse temporal cortex | 2.35 (0.14) | 2.23 (0.15) | 0.006 |
|  | Middle temporal gyrus | 2.68 (0.12) | 2.60 (0.09) | 0.007 |
|  | Insula | 2.85 (0.13) | 2.73 (0.13) | 0.007 |
|  | Pars opercularis | 2.45 (0.09) | 2.38 (0.10) | 0.022 |
|  | Banks of superior temporal sulcus | 2.38 (0.08) | 2.32 (0.10) | 0.026 |
|  | Paracentral lobule | 2.34 (0.13) | 2.26 (0.12) | 0.037 |
| Thickness, right | Pars triangularis | 2.29 (0.12) | 2.19 (0.08) | 0.007 |
|  | Middle temporal gyrus | 2.73 (0.10) | 2.65 (0.11) | 0.010 |
|  | Paracentral lobule | 2.39 (0.14) | 2.32 (0.12) | 0.010 |
|  | Postcentral gyrus | 2.07 (0.12) | 1.97 (0.12) | 0.011 |
|  | Transverse temporal cortex | 2.36 (0.14) | 2.24 (0.18) | 0.016 |
|  | Superior temporal gyrus | 2.65 (0.13) | 2.56 (0.14) | 0.028 |
|  | Lateral occipital cortex | 2.23 (0.09) | 2.18 (0.08) | 0.048 |
| Surface area, left | Superior temporal gyrus | 0.0026 (0.0002) | 0.0028 (0.0002) | 0.021 |
|  | Fusiform gyrus | 0.0021 (0.0002) | 0.0022 (0.0002) | 0.031 |
|  | Paracentral lobule | 0.0009 (0.0001) | 0.0010 (0.0001) | 0.041 |
| Surface area, right | Precuneus cortex | 0.0027 (0.0003) | 0.0029 (0.0003) | 0.031 |
|  | Paracentral lobule | 0.0010 (0.0001) | 0.0011 (0.0001) | 0.048 |
| Volume, right | Fusiform gyrus | 0.0060 (0.0008) | 0.0065 (0.0009) | 0.025 |
| Deformation, left | Pallidum | 0.92 (0.05) | 0.88 (0.05) | 0.020 |
|  | Insula | 1.00 (0.05) | 1.04 (0.04) | 0.031 |
| Deformation, right | Isthmus of cingulate cortex | 1.01 (0.06) | 0.97 (0.05) | 0.007 |
|  | Putamen | 0.92 (0.05) | 0.88 (0.04) | 0.026 |
|  | Pallidum | 0.92 (0.05) | 0.89 (0.04) | 0.033 |

iRBD: idiopathic rapid eye movement sleep behavior disorder. MCI: mild cognitive impairment. Surface area and volume values were normalized using the estimated total intracranial volume. Red: Thinning, reduced surface area or volume, or tissue deformation value.

**Supplementary Figure 1.**
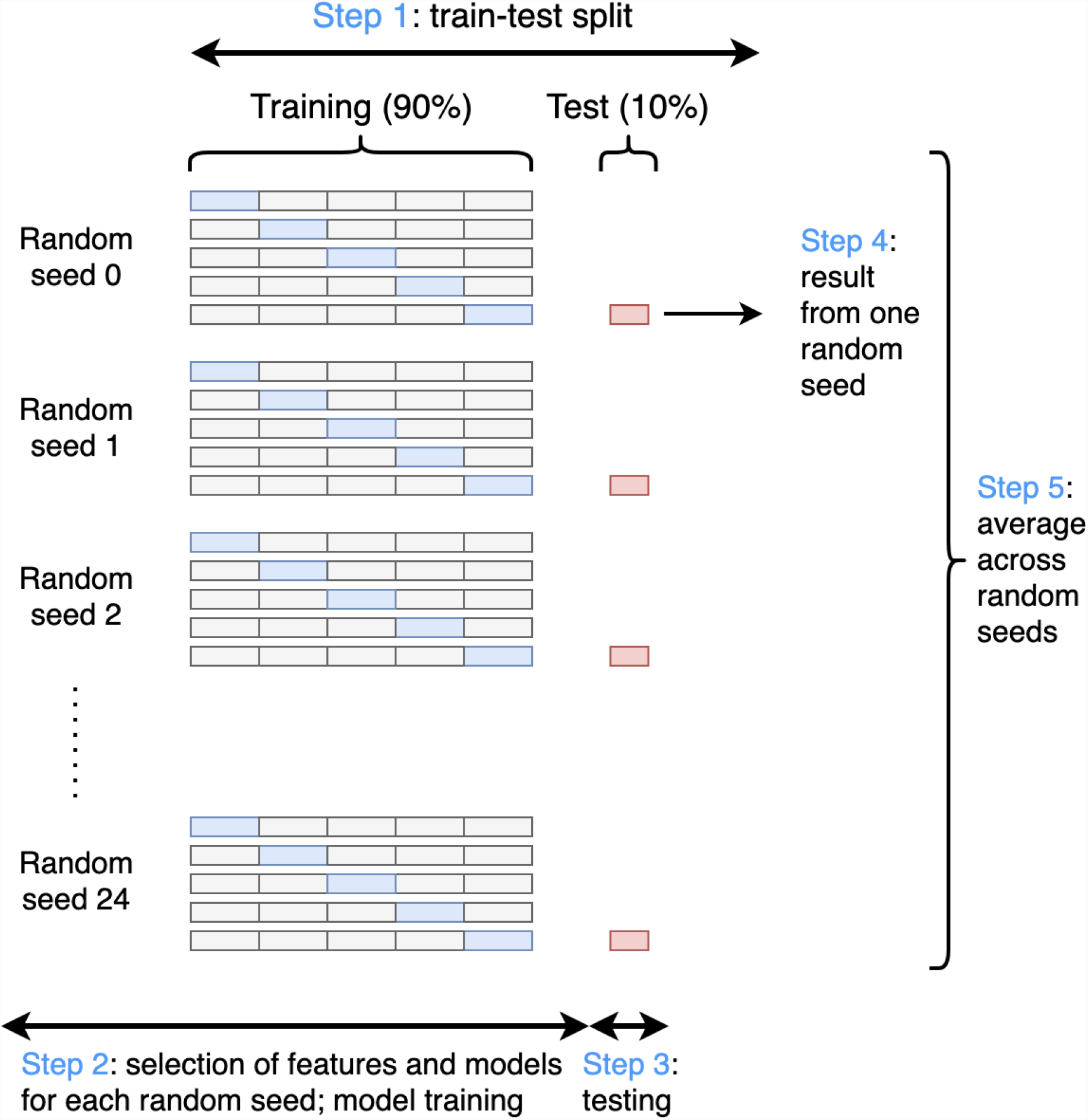
A diagram demonstrating all steps of the proposed analysis, including random train-test split, feature and model selection, model training and testing, and evaluation of model performance.

## Notes

**Conflict of interest:** The authors declare that there is no conflict of interest.

### Competing Interest Statement

The authors have declared no competing interest.

### Author Declarations

All participants were part of research protocols approved by local ethics committees (CIUSSS-NÎM-HSCM) and CIUSSS du Centre-Sud-de-l'Île-de-Montréal-Comité d'éthique de la recherche vieillissement-neuroimagerie, Montreal, Canada) and provided written informed consent.

### Summary of Updates

Figures 2 and 3 revised. Multiple paragraphs revised.

